# HiFi sequencing accurately identifies clinically relevant variants in paralogous genes

**DOI:** 10.1101/2025.10.29.25339045

**Authors:** Bart van der Sanden, Christian Betz, Katharina Herzog, Esther Schamschula, Katharina Wimmer, Inga Vater, Saranya Balachandran, Xiao Chen, Jordi Corominas Galbany, Raoul Timmermans, Ronny Derks, HiFi Solves EMEA Consortium, Malte Spielmann, Michael A. Eberle, Christian Gilissen, Lisenka E.L.M. Vissers, Johannes Zschocke, Hanno J. Bolz, Alexander Hoischen

## Abstract

Short-read sequencing (SRS) methods have improved the detection of small genetic variants but remain limited in highly homologous genomic regions, such as segmental duplications with gene-pseudogene pairs. These paralogous regions often require complex, locus-specific assays for accurate analysis. Long-read genome sequencing (lrGS) technologies, such as PacBio HiFi sequencing, can span these regions but still face challenges in variant calling due to alignment ambiguities. Here, we evaluated PacBio HiFi lrGS combined with Paraphase, a dedicated haplotype-based variant caller, in 86 individuals with 125 known clinically relevant variants across 11 paralogous loci. Standard HiFi variant callers detected 95/125 variants, while the remaining 30 variants were only identified by Paraphase. Together, the standard variant callers and Paraphase detected all known variants, including SNVs, InDels, CNVs, SVs, and gene conversions. In addition, lrGS allowed accurate phasing and gene-pseudogene copy number detection. We demonstrate that PacBio HiFi lrGS, particularly when integrated with Paraphase, enables comprehensive variant detection in previously difficult-to-assess genomic regions. These results also suggest that lrGS is ready for a wider implementation, possibly as a first-tier diagnostic approach for individuals with suspected variants in these paralogous regions.

## Main text

Short-read sequencing (SRS) methods, such as gene panel, exome, and genome sequencing, have significantly increased the diagnostic yield with regard to short genetic variants, including substitutions and small InDels^1^. However, independent of variant types, several regions in the genome remain difficult to interrogate by standard SRS reads^2-4^. These difficult regions are also often referred to as “dark” or “camouflaged” regions^3^. Many of these camouflaged regions are regions with high sequence homology that arise from segmental duplications or other duplicated genomic regions^5^. The unique sequences within these duplicated regions cannot be fully spanned by short reads, preventing accurate distinction between the duplicated copies and thereby hindering comprehensive and reliable variant calling in these regions^3^. Several disease-associated genes are located in segmental duplications, and many exist as functional gene-pseudogene paralog pairs, several even with multiple pseudogenes. Often, genetic variation cannot be reliably detected in these regions, *e*.*g*. those containing *OTOA/OTOAP1, CYP21A2/CYP21A1P, PMS2/PMS2CL* and *STRC/STRCP1*. The standard analysis of genetic variation in these medically relevant genes includes targeted testing, which is sometimes only available for a small number of known specific variants^6,7^. For each locus, these tests have to be customized, which is a laborious process, as multiple different tests such as transcript analysis, allele-specific PCR, and MLPA may be needed in addition to SRS.

Long-read genome sequencing (lrGS) technologies, *e*.*g*. PacBio HiFi and Oxford Nanopore Technologies (ONT), now enable the detection of all variant types and interrogation of even the most difficult regions of the entire genome, including the regions with very high sequence homology^3,8,9^. However, even among these highly accurate long-read assemblies, the segmental duplication regions are still enriched for assembly errors^10^. To overcome these assembly errors, Chen *et al*. developed Paraphase^11^, a tool to identify haplotypes of genes and their paralogs. Originally aiming at the *SMN1*/*SMN2* region, it was subsequently optimized for 159 other clinically relevant segmental duplication regions across the entire genome^7^. Paraphase enabled the accurate detection of genetic variation in these homologous regions that remained difficult to genotype before.

Here, we assessed the technical validity of PacBio HiFi genome sequencing in combination with Paraphase to detect genetic variants in clinically relevant gene/pseudo-gene combinations. To this end, we performed PacBio HiFi lrGS for 86 individuals with one or more known clinically relevant genetic variants in 11 genomic regions with high sequence homology. We evaluated the capability of HiFi lrGS to accurately detect all classes of variants, including single nucleotide variants (SNVs), insertions/deletions (InDels), copy number variants (CNVs), structural variants (SVs), and gene conversions underlying these variants within 11 paralogous regions. For all samples, we ran the standard HiFi genome sequencing and variant analysis pipeline (**Supplementary Methods**). Four regions (*ABCC6, PKD1, RPGR* and *SHOX*) had relatively low sequence similarity (<99%) between the functional protein-coding genes and paralogs/pseudogenes, which should allow standard HiFi analysis to distinguish the homologous copies and to call variants in these regions. The seven remaining regions (*CYP21A2, IKBKG, OTOA, PMS2, SMN1*/*SMN2, STRC* and *TNXB*) had higher sequence similarity between paralogs and were thus included in Paraphase for analysis. This not only allowed us to compare the HiFi data to current standard methods, including Sanger sequencing, MLPA, CNV microarrays, and short-read sequencing, but also to directly compare the standard lrGS analysis pipeline with Paraphase. We then assessed whether PacBio HiFi sequencing can now be used to detect variation in these extremely difficult genomic regions.

In total, the retrospectively selected samples contained 125 known clinically relevant variants in the 11 different homologous regions (**Table S1**). These variants included 58 SNVs, 26 InDels and 41 SVs (**Figure 1A**). In total, 134 standard of care (SoC) tests were performed for these 86 samples, ranging from 1-3 tests per sample. SoC included Sanger sequencing, exome sequencing, short-read panel sequencing, short-read amplicon sequencing, MLPA, long-read amplicon sequencing, single-molecule molecular inversion probes (smMIPs), allele-specific PCR, and transcript analysis (**Figure 1B**). In standard of care, all CNVs and SVs were detected using MLPA or long-read amplicon sequencing, but none by short-read sequencing. This was primarily due to the poor mapping quality of the short reads in these homologous regions (**Figure 2**) and the use of exome sequencing rather than whole-genome sequencing for these samples. While a short-read genome already offers benefits over exome sequencing, long-read genomes provide a significant advantage over SRS for SV calling, as they can interrogate regions of the genome that are inaccessible to SRS and make them available to variant callers^8,12^.

**Figure 1:**
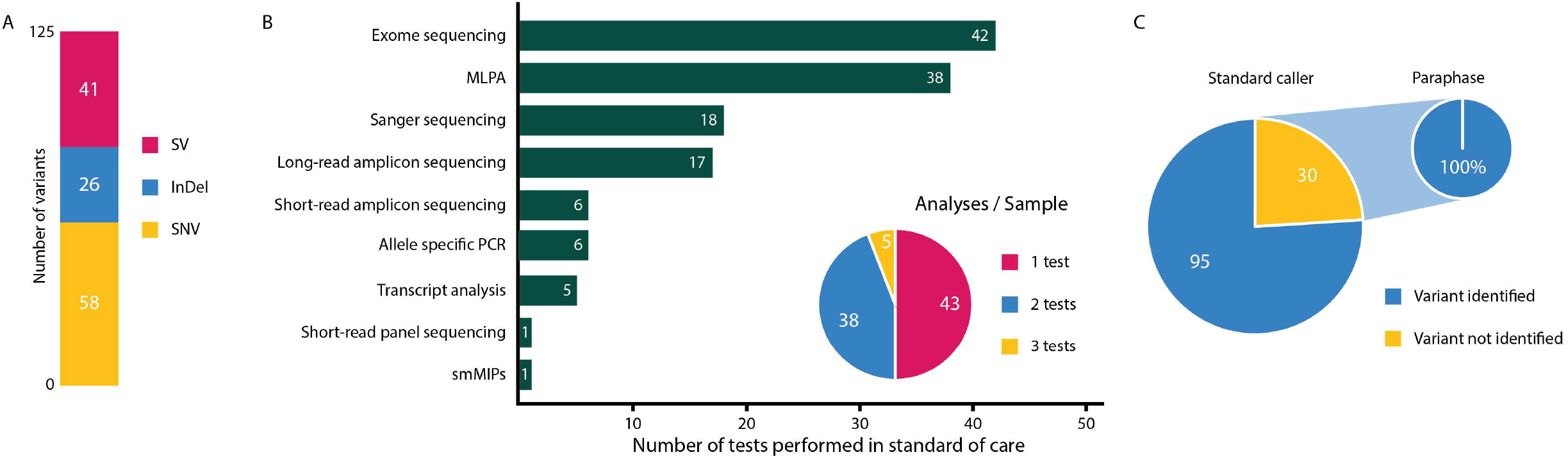
Variants, standard of care and lrGS results. (A) Bar graph representing the number of each variant type included in the 125 known clinically relevant variants. (B) Overview of the number of standard of care tests that were used to identify the variants. (C) lrGS results with the large pie chart representing the findings from the standard callers. Of the 125 variants, 30 were not identified by the standard callers, but all 30 were later recovered by Paraphase, as shown in the small pie chart.

**Figure 2:**
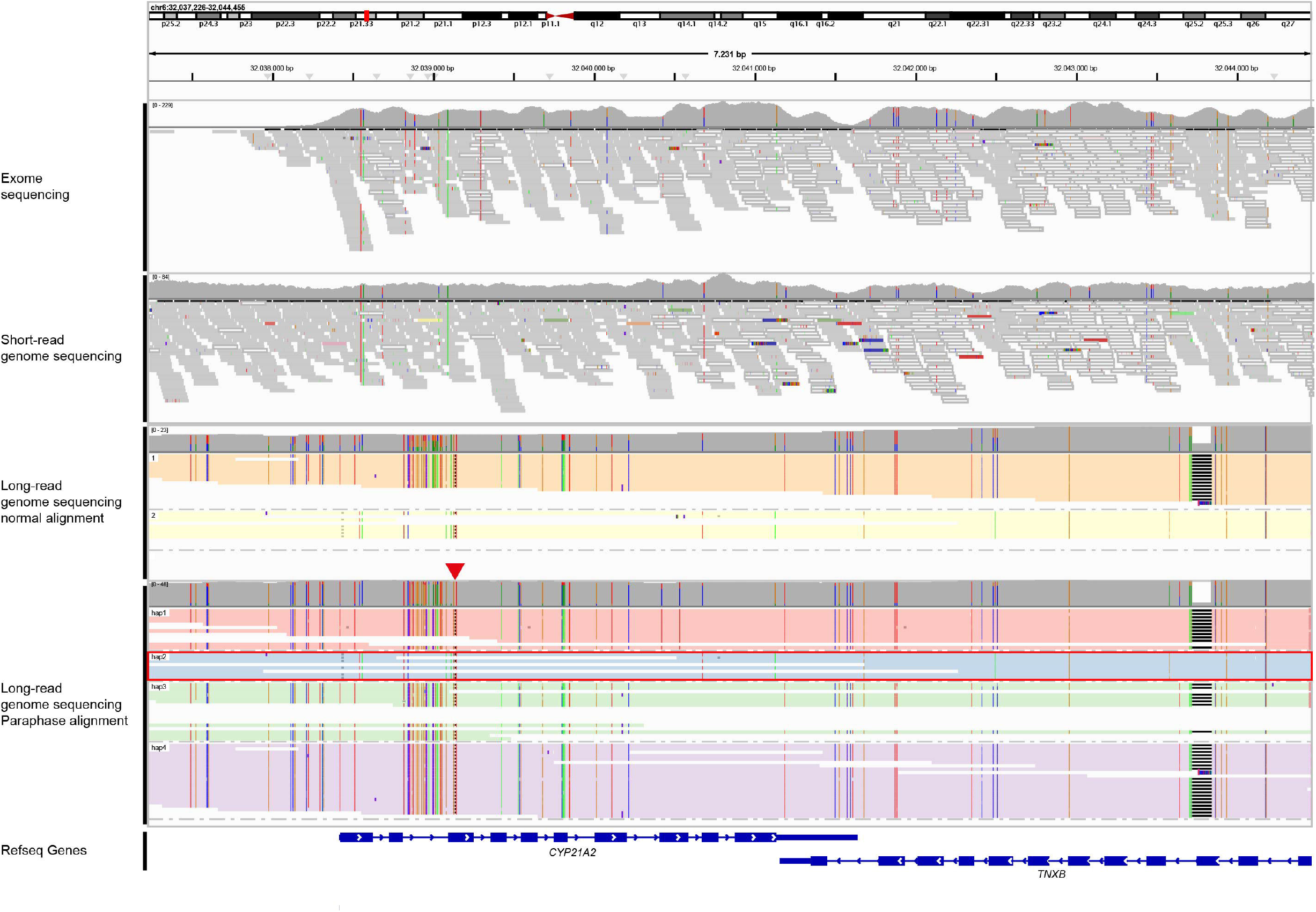
Read mapping comparison between exome, short-read genome and long-read genome sequencing for the paralogous CYP21A2 region in sample S10. Exome and short-read genome data are representative datasets and are not from the same individual. For lrGS, both the normal alignment and Paraphase alignment are presented. The reads in the long-read genome sequencing alignments are colored by haplotype. Gray reads are reads with sufficient mapping quality while white reads indicate reads with mapping quality 0. These white reads typically appear in the exome and short-read genome sequencing at the parts of this region with highest sequence similarity between genes and paralogs. In the Paraphase alignment, reads were grouped by the phase assigned by Paraphase (hap1-4). Hap1, hap3 and hap4 were considered pseudogene copies, while hap2 (colored light blue and highlighted by the red rectangle) was considered a functional gene copy. For this specific sample (S10), two clinically relevant variants were reported before. The first variant was an indel event that is also indicated by the red arrow head in the Paraphase alignment. The other event was a deletion of one copy of CYP21A2, which was also identified by lrGS by detecting a reduced functional gene copy number of one.

We performed HiFi long-read sequencing for these 86 samples according to the manufacturer’s protocol. In short, we used a single SMRT cell per sample to generate an expected coverage of 30X. Variant calling was performed using the standard variant callers by PacBio, including DeepVariant, pbsv and HiFiCNV. In addition, we used Paraphase. We obtained a median HiFi output of 92.32 Gb, corresponding to a median genome-wide coverage of 28.6X, with a median average read length of 15.5 kb (**Table S2**). Of the 125 known variants, 95, in nine different genomic loci, were identified by the variant type-specific standard variant calling tool (**Figure 1C; Table S1**). This included 58 SNVs, 25 InDels and 12 SVs. All 95 variants were detected from the variant calling files and manual inspection was not necessary. The remaining 30 variants were not detected and were also not identified after manual inspection. These included one InDel and 29 SVs in *CYP21A2, IKBKG, OTOA, PMS2* / *PMS2CL, SMN1* / *SMN2*, and *STRC*. All 30 variants were identified by Paraphase (**Figure 1C**). This was mainly due to the clear haplotype separation of the pseudogene and functional gene copies by Paraphase (**Figure 3A**). Jointly, we were able to identify all 125 clinically relevant variants affecting the 11 different loci in 86 samples (**Figure 1C; Figure S1; Table S1**).

**Figure 3:**
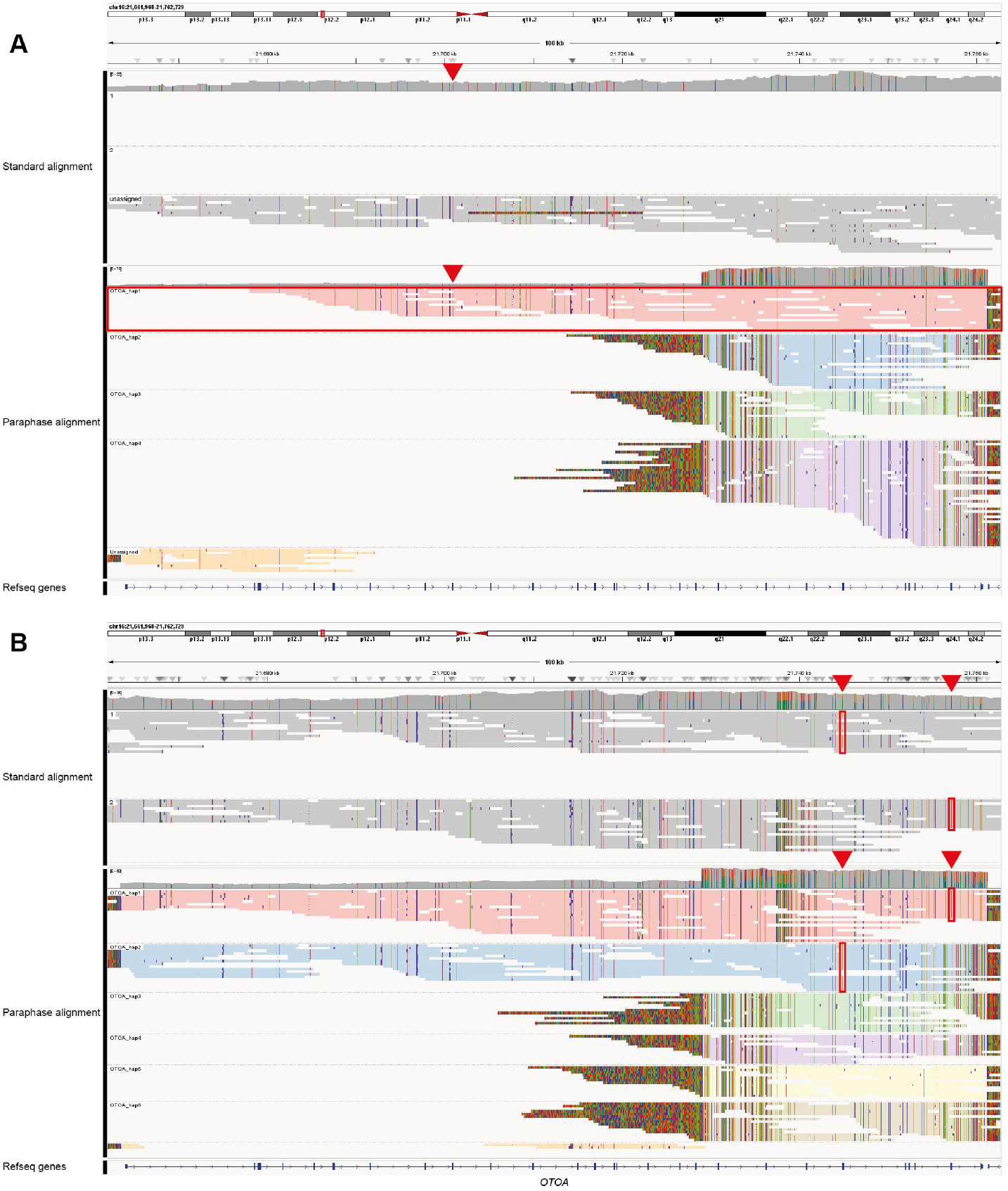
IGV comparison of the normal alignment versus the Paraphase alignment. **A)** For sample S26, a deletion and an SNV were reported. The top panel represents the standard alignment where reads were grouped by phase. Phasing was unsuccessful and therefore all reads ended up in the unassigned box. The bottom panel represents the Paraphase alignment and reads were grouped and colored by phase. Because Paraphase distinguishes between functional gene copies and pseudogene copies, more than two haplotypes were assigned. In this case there were 4 haplotypes (marked OTOA_hap1 - OTOA_hap4). OTOA_hap2 - OTOA_hap4 were the pseudogene copies, while OTOA_hap1 was the functional gene copy. This means there was only one functional gene copy, corresponding with the finding in standard of care. The SNV was also identified and is indicated by the red arrowhead. **B)** For sample S24, three events were detected, two SNVs and one indel. The top panel again shows the normal alignment and the bottom panel the Paraphase alignment. In both panels, reads were grouped by phase. In the normal alignment phasing was successful and all reads were assigned to either haplotype 1 or haplotype 2. In the Paraphase alignment there were six haplotypes, of which two functional gene copies (OTOA_hap1 and OTOA_hap2) and four pseudogene copies (OTOA_hap3 - OTOA_hap6). The indel event was located outside the homology region and is not visible in this figure. The two SNVs were assigned to the functional gene copies (indicated by the red arrowheads and red boxes). By using this accurate phasing strategy we were able to identify that both variants were on a different haplotype and therefore in trans.

Paraphase v.3.0.0 supported seven of the 11 loci as these had long (>10kb) regions of high (>99%) sequence similarity between genes and paralogs/pseudogenes (Table S1)^7^. These seven loci contained 79 of the 125 variants in this cohort (**Supplementary Table S3**). Of these 79 variants in a Paraphase-supported locus, four were not identified by the Paraphase software and one required an additional bioinformatic analysis to identify the event (**Figure S1; Table S1**). Three of the four missed variants were not identified because the version of Paraphase that was used (v3.0.0) only supported variant calling in the homology region of *TNXB*. Our dataset included four variants in *TNXB*, one of which was located in the homology region and this variant was also identified by Paraphase. The other three variants were in the non-homologous region and were called by the standard variant caller only. Of note, Paraphase versions 3.2.0 or later, support the entire *TNXB* gene including the non-homologous portion of the gene. The remaining event that was not identified by Paraphase, was a one base pair insertion and the haplotype carrying the expected variant was only present in two of the reads, which is below the depth threshold that Paraphase is designed to work with (**Figure S2**). Finally, in sample S42, we needed to run Sawfish^13^, a separate structural variant caller, to detect the ∼2 kb insertion event. Currently, variant calling in Paraphase is performed using a simple pileup-based approach, which requires that most reads supporting a given haplotype contain the variant in the alignment. As a result, Paraphase is primarily suited for calling small variants. In the case of this larger SV, several reads were soft-clipped at the insertion site, and variability in insertion length introduced further complexity (**Figure S3**). Consequently, there was insufficient evidence for Paraphase to detect the event. In the future, updates to Paraphase may eliminate the need for this additional bioinformatic step.

One of the benefits of lrGS over short-read exome and genome sequencing is its more uniform and high quality coverage for the paralogous regions (**Figure 2**). However, coverage is also a sensitive factor for Paraphase, as reduced coverage causes the steepest decline in the discovery rate of CNVs and SNVs that require Paraphase, across all variant types^14^. This may require a slightly higher coverage for samples with suspected variants in paralogous regions. An additional benefit of lrGS is its ability to determine the copy number of the protein-coding gene and pseudogene (**Figure 2**) and that it can identify potential gene conversions or micro-conversions. For example, in sample S19, with a reported duplication in *CYP21A2*, we were able to determine that the *CYP21A2*/*CYP21A1P* copy number was normal and that each allele had one protein-coding gene copy and one pseudogene copy (**Figure S4**). A part of one pseudogene copy showed gene conversion with the protein-coding gene, a well-known process for *CYP21A2*^15^, which has also likely caused the MLPA to detect a duplication as there was more material of the protein-coding gene because of the micro-conversion. This method not only worked for *CYP21A2*, but also for the other genes with reported copy number changes. In sample S26, an SNV and a deletion were reported. The SNV was identified by DeepVariant and Paraphase, however, the deletion was only detected by Paraphase and not by the standard structural variant caller pbsv. Inspection of the Paraphase alignment is very intuitive for identifying such events, as it clearly distinguishes the protein-coding gene copy from the pseudogene copies, indicating a heterozygous deletion of *OTOA* (**Figure 3A**). Phasing the reads in the normal alignment was unsuccessful, as all reads were placed in the unassigned group. Retrospectively, this was likely due to the deletion, but taking failed phasing as a measure for a deletion or other structural variant is less intuitive than using the Paraphase alignment. In addition to determining gene copy number, lrGS can be used to determine whether variants occurred in *cis* or in *trans*. Correctly phasing, or assigning variants to a specific haplotype, can change the interpretation of a patient’s genotype from pathogenic to benign and *vice versa*, a process very important for assessing inheritance patterns and recurrence risks for family members. For example, sample S24 had three known clinically-relevant variants in *OTOA*. Two of these were a SNV and the other was a one base pair deletion. All three variants were identified by the standard small variant caller DeepVariant, as well as Paraphase, but lrGS now allowed us to determine that the two SNVs were located in the protein-coding gene *OTOA*. Additionally, by phasing the SNVs, we determined that the variants were in *trans*, meaning that both alleles were affected and these events are likely disease causing (**Figure 3B**). Finally, by integrating both the standard variant callers and Paraphase in the HiFi lrGS interpretation workflow, all phased variants and gene copy numbers can be identified in a single genetic test. With further improvements of detection tools, possibly in combination with the integration of a *de novo* assembly, we expect that even the most difficult variants can be detected automatically in the future.

Variant detection and interpretation in clinically relevant paralogous sequences belong to the most challenging tasks in human genetics. The ability to analyze even these highly homologous loci of the human genome using lrGS provides great promise for clinical care. In this study, we show that all 125 clinically relevant variants, that were known to be present in 11 different paralogous regions in 86 different patients, were identified by HiFi lrGS. For 30/125 (24.0%) of the variants, Paraphase, a dedicated HiFi-based variant caller for these highly similar paralogous genes, was required because the variant calling results from the respective standard variant caller were not sufficient to identify the known event. Paraphase (v3.0.0) was developed for 160 of these paralogous regions with >99% sequence homology, including 316 genes, and has shown copy number population diversity across many of those loci^7^. Our approach of combining the general variant callers with Paraphase was successful for the full range of variant types *i*.*e*., SNVs, InDels, SVs, CNVs, and gene conversions including heterozygous and homozygous events in autosomal recessive and autosomal dominant genes. Although this is a proof-of-concept study based on positive controls with known pathogenic variants, our results indicate that HiFi lrGS is highly suitable for routine diagnostic application that also covers such difficult-to-detect events.

In an earlier study, 8/57 (14.0%) variants in the same 11 loci that we assessed in this study, were not detected by short-read genome sequencing^4^. Notably, all eight variants were located in the seven genes also supported by Paraphase. In that same study by Schobers *et al*., 14 additional variants in homologous regions in three different patients were not detected by short-read genome sequencing. Two of these samples were later included in another study assessing HiFi lrGS for difficult-to-detect, clinically relevant variants^14^. All variants in these two samples were identified by Paraphase, confirming our finding that long-read sequencing has improved variant calling for these very homologous regions in the genome.

Beyond the seven loci supported by Paraphase, we included four other clinically relevant paralogous genes for which Paraphase was not required. Our results highlight that just the general variant caller was already sufficient to identify all the clinically relevant variants in these genes. This indicates that developing Paraphase for these loci may not be necessary, since the relatively low (<99%) or short (<10kb) sequence similarity between genes and paralogs/pseudogenes makes it possible for standard HiFi analysis to call variants accurately.

Our current results demonstrate the abilities of HiFi genome sequencing to identify clinically-relevant variants in homologous sequences. In addition to variant calling in these homologous regions, lrGS can be used to accurately detect other difficult-to-detect, clinically relevant variants, such as repeat expansions, complex rearrangements, and differential methylation, such as imprinting and skewed X-inactivation^14^. lrGS has also been shown to yield additional genetic diagnoses in cohorts of previously undiagnosed patients^16^. Conceptually, this study also supports that lrGS starts enabling to move from a variant centric view in human genetics to a allelic and haplotype-resolved view. Together this suggests a potential role for HiFi long-read sequencing as first-tier genetic test. However, further prospective studies evaluating both the clinical utility and cost-effectiveness of lrGS alongside the standard of care are warranted.

## Data Availability

All data produced in the present study are available upon reasonable request to the authors.

## Acknowledgements

We thank our colleagues from the clinical and diagnostic division of the Klinisch Genetisch Centrum Nijmegen as well as Radboud Genome Technology Center, particularly T. Hofste, A. van Ouden and Dr. J. Weiss, and the Netherlands X-omics Initiative NWO (project 184.034.019), for technical and financial support. AH was supported by a ZonMW (The Netherlands Organization for Health Research and Development) Vici grant (No. 09150182310053). The aims of this study contribute to the Solve-RD project (to AH, CG and LELMV), which has received funding from the European Union’s Horizon 2020 research and innovation program under grant agreement No 779257. The aims of this study contribute to the ERDERA project (to BvdS, AH, CG and LELMV). ERDERA has received funding from the European Union’s Horizon Europe research and innovation programme under grant agreement N°101156595. Views and opinions expressed are those of the author(s) only and do not necessarily reflect those of the European Union or any other granting authority, who cannot be held responsible for them.

## References

1. Karczewski, K.J., Francioli, L.C., Tiao, G., Cummings, B.B., Alföldi, J., Wang, Q., Collins, R.L., Laricchia, K.M., Ganna, A., Birnbaum, D.P., et al. (2020). The mutational constraint spectrum quantified from variation in 141,456 humans. Nature 581, 434–443. 10.1038/s41586-020-2308-7.

2. Chaisson, M.J.P., Sanders, A.D., Zhao, X., Malhotra, A., Porubsky, D., Rausch, T., Gardner, E.J., Rodriguez, O.L., Guo, L., Collins, R.L., et al. (2019). Multi-platform discovery of haplotype-resolved structural variation in human genomes. Nature Communications 10, 1784. 10.1038/s41467-018-08148-z.

3. Ebbert, M.T.W., Jensen, T.D., Jansen-West, K., Sens, J.P., Reddy, J.S., Ridge, P.G., Kauwe, J.S.K., Belzil, V., Pregent, L., Carrasquillo, M.M., et al. (2019). Systematic analysis of dark and camouflaged genes reveals disease-relevant genes hiding in plain sight. Genome Biology 20, 97. 10.1186/s13059-019-1707-2.

4. Schobers, G., Derks, R., den Ouden, A., Swinkels, H., van Reeuwijk, J., Bosgoed, E., Lugtenberg, D., Sun, S.M., Corominas Galbany, J., Weiss, M., et al. (2024). Genome sequencing as a generic diagnostic strategy for rare disease. Genome Med 16, 32. 10.1186/s13073-024-01301-y.

5. Bailey, J.A., Gu, Z., Clark, R.A., Reinert, K., Samonte, R.V., Schwartz, S., Adams, M.D., Myers, E.W., Li, P.W., and Eichler, E.E. (2002). Recent segmental duplications in the human genome. Science 297, 1003–1007. 10.1126/science.1072047.

6. Haer-Wigman, L., den Ouden, A., van Genderen, M.M., Kroes, H.Y., Verheij, J., Smailhodzic, D., Hoekstra, A.S., Vijzelaar, R., Blom, J., Derks, R., et al. (2022). Diagnostic analysis of the highly complex OPN1LW/OPN1MW gene cluster using long-read sequencing and MLPA. NPJ Genom Med 7, 65. 10.1038/s41525-022-00334-9.

7. Chen, X., Baker, D., Dolzhenko, E., Devaney, J.M., Noya, J., Berlyoung, A.S., Brandon, R., Hruska, K.S., Lochovsky, L., Kruszka, P., et al. (2025). Genome-wide profiling of highly similar paralogous genes using HiFi sequencing. Nature Communications 16, 2340. 10.1038/s41467-025-57505-2.

8. Kucuk, E., van der Sanden, B., O’Gorman, L., Kwint, M., Derks, R., Wenger, A.M., Lambert, C., Chakraborty, S., Baybayan, P., Rowell, W.J., et al. (2023). Comprehensive de novo mutation discovery with HiFi long-read sequencing. Genome Med 15, 34. 10.1186/s13073-023-01183-6.

9. Nurk, S., Koren, S., Rhie, A., Rautiainen, M., Bzikadze, A.V., Mikheenko, A., Vollger, M.R., Altemose, N., Uralsky, L., Gershman, A., et al. (2022). The complete sequence of a human genome. Science 376, 44–53. doi:10.1126/science.abj6987.

10. Vollger, M.R., Dishuck, P.C., Harvey, W.T., DeWitt, W.S., Guitart, X., Goldberg, M.E., Rozanski, A.N., Lucas, J., Asri, M., Munson, K.M., et al. (2023). Increased mutation and gene conversion within human segmental duplications. Nature 617, 325–334. 10.1038/s41586-023-05895-y.

11. Chen, X., Harting, J., Farrow, E., Thiffault, I., Kasperaviciute, D., Hoischen, A., Gilissen, C., Pastinen, T., and Eberle, M.A. (2023). Comprehensive SMN1 and SMN2 profiling for spinal muscular atrophy analysis using long-read PacBio HiFi sequencing. Am J Hum Genet 110, 240–250. 10.1016/j.ajhg.2023.01.001.

12. Logsdon, G.A., Vollger, M.R., and Eichler, E.E. (2020). Long-read human genome sequencing and its applications. Nat Rev Genet 21, 597–614. 10.1038/s41576-020-0236-x.

13. Saunders, C.T., Holt, J.M., Baker, D.N., Lake, J.A., Belyeu, J.R., Kronenberg, Z., Rowell, W.J., and Eberle, M.A. (2025). Sawfish: improving long-read structural variant discovery and genotyping with local haplotype modeling. Bioinformatics 41. 10.1093/bioinformatics/btaf136.

14. Höps, W., Weiss, M.M., Derks, R., Galbany, J.C., Ouden, A.D., van den Heuvel, S., Timmermans, R., Smits, J., Mokveld, T., Dolzhenko, E., et al. (2025). HiFi long-read genomes for difficult-to-detect, clinically relevant variants. Am J Hum Genet 112, 450–456. 10.1016/j.ajhg.2024.12.013.

15. Carrozza, C., Foca, L., De Paolis, E., and Concolino, P. (2021). Genes and Pseudogenes: Complexity of the RCCX Locus and Disease. Front Endocrinol (Lausanne) 12, 709758. 10.3389/fendo.2021.709758.

16. Steyaert, W., Sagath, L., Demidov, G., Yépez, V.A., Esteve-Codina, A., Gagneur, J., Ellwanger, K., Derks, R., Weiss, M., den Ouden, A., et al. (2025). Unraveling undiagnosed rare disease cases by HiFi long-read genome sequencing. Genome Res 35, 755–768. 10.1101/gr.279414.124.

