## Supplementary Materials for "HiFi sequencing accurately identifies clinically relevant variants in paralogous genes"

* Full list of consortium members shown in supplement.

[#] Corresponding author:
Alexander Hoischen, PhD
Department of Human Genetics, Radboudumc
Geert Grooteplein 10, 6500HB Nijmegen, The Netherlands


### Supplementary Figures

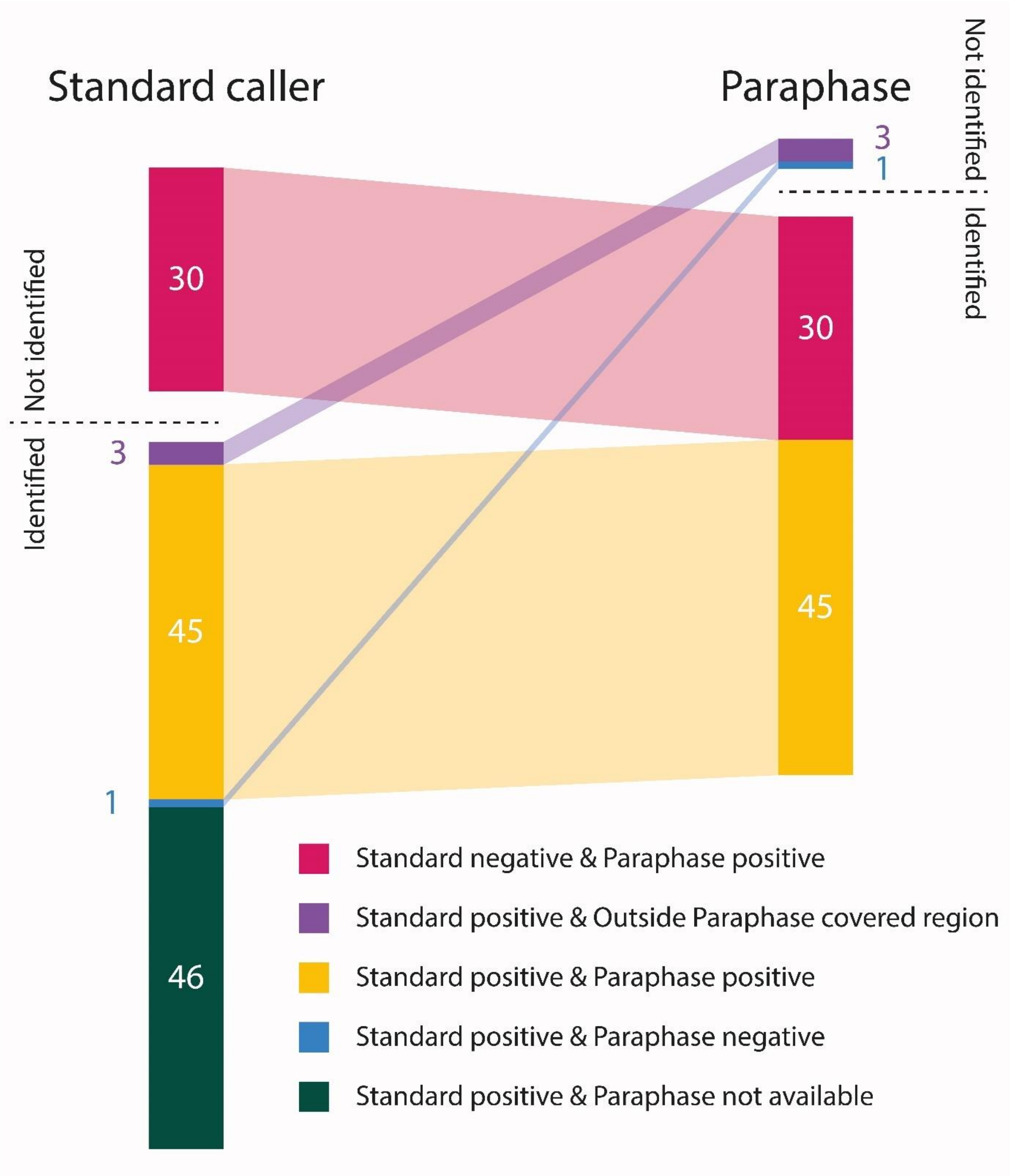


**Figure S1: Overview of the number of variants identified by the standard callers and Paraphase.**

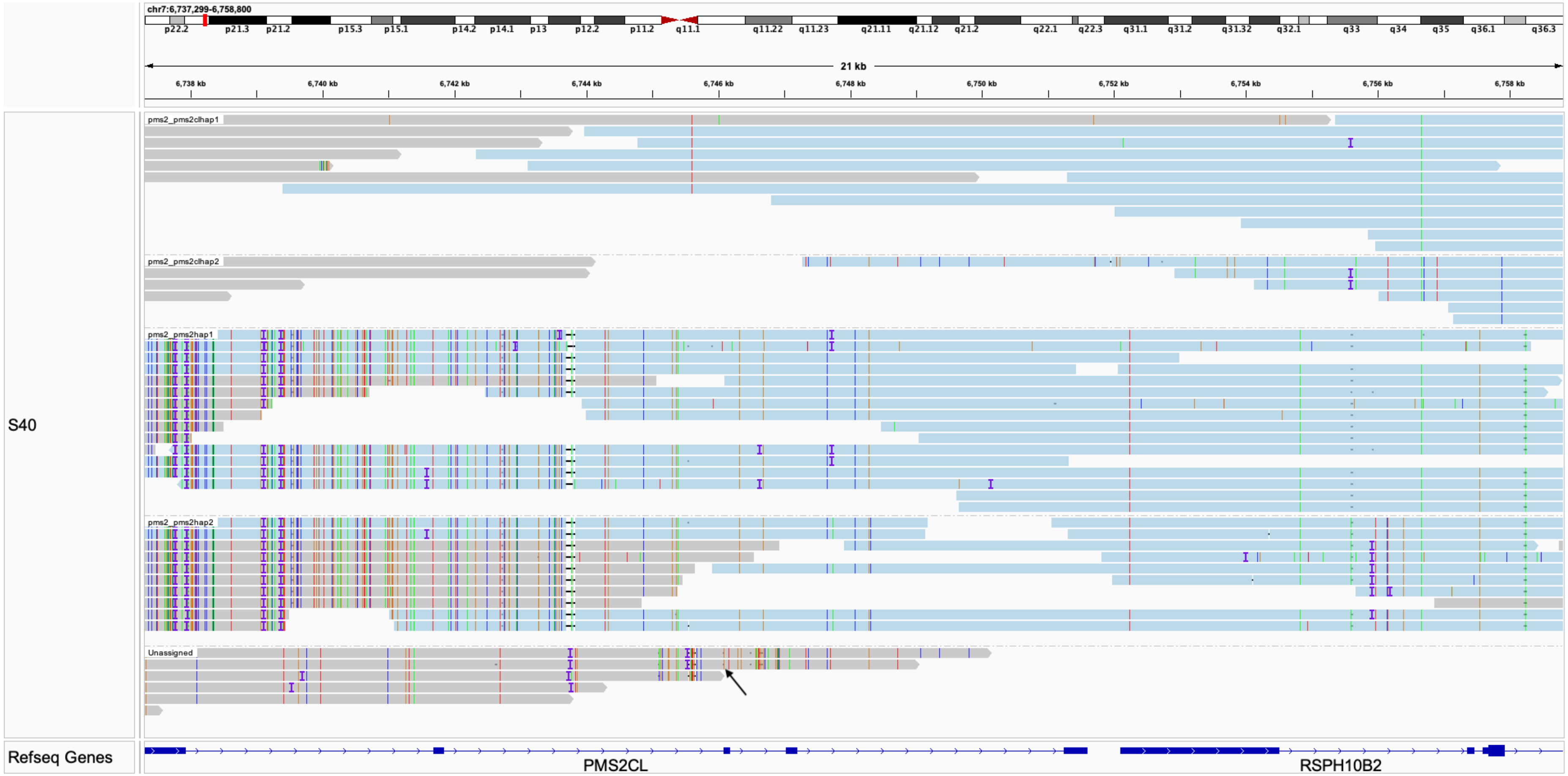


**Figure S2: A small variant in *PMS2CL* missed by Paraphase.**

Paraphase-phased haplotypes are shown against the *PMS2CL* reference. Four haplotypes were phased by Paraphase, two *PMS2CL* copies (top) and two *PMS2* copies (bottom). The second *PMS2CL* haplotype was only partially phased due to low coverage. The missed small variant (marked by the black arrow) was present in only two reads, which were not incorporated into the second *PMS2CL* haplotype and were labeled “Unassigned”.

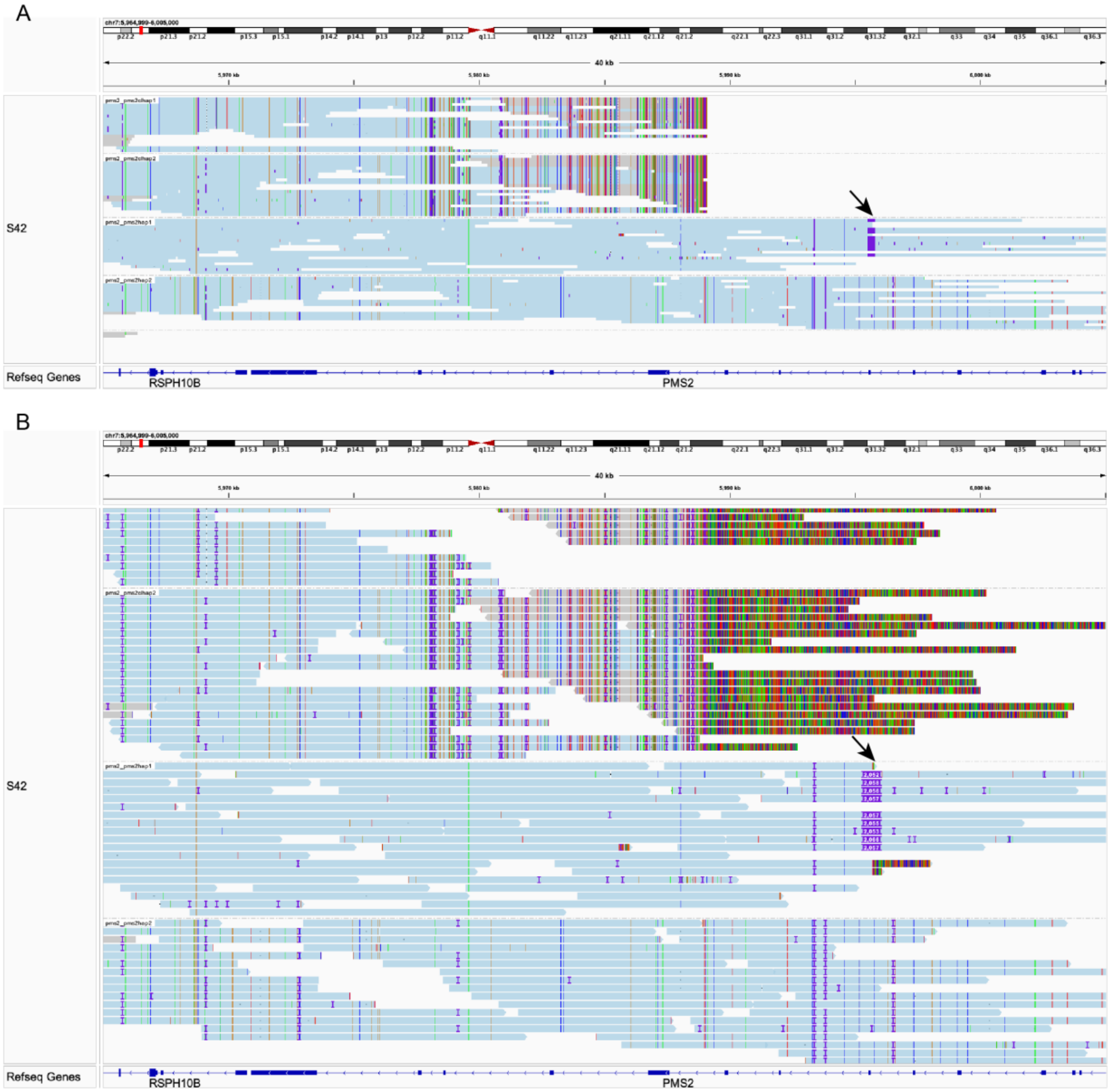


**Figure S3: A 2057bp insertion in *PMS2* missed by Paraphase.**

(A) Four haplotypes were phased by Paraphase, two *PMS2CL* copies (top) and two *PMS2* copies (bottom). The *PMS2CL* copies are shorter as the homology region only spans the last five exons of *PMS2*. All reads from the third haplotype (a *PMS2* copy) carry the insertion (black arrow). The insertion was not called by Paraphase because Paraphase currently does not have a built-in caller for large SVs. This SV was called after running Sawfish (Version 1.0.2) on the Paraphase output BAM. With future incorporation of an SV caller into the Paraphase workflow, large SVs like this can be called by Paraphase. (B) Expanded view of A, showing soft-clipped bases and the size of the insertion on each read.

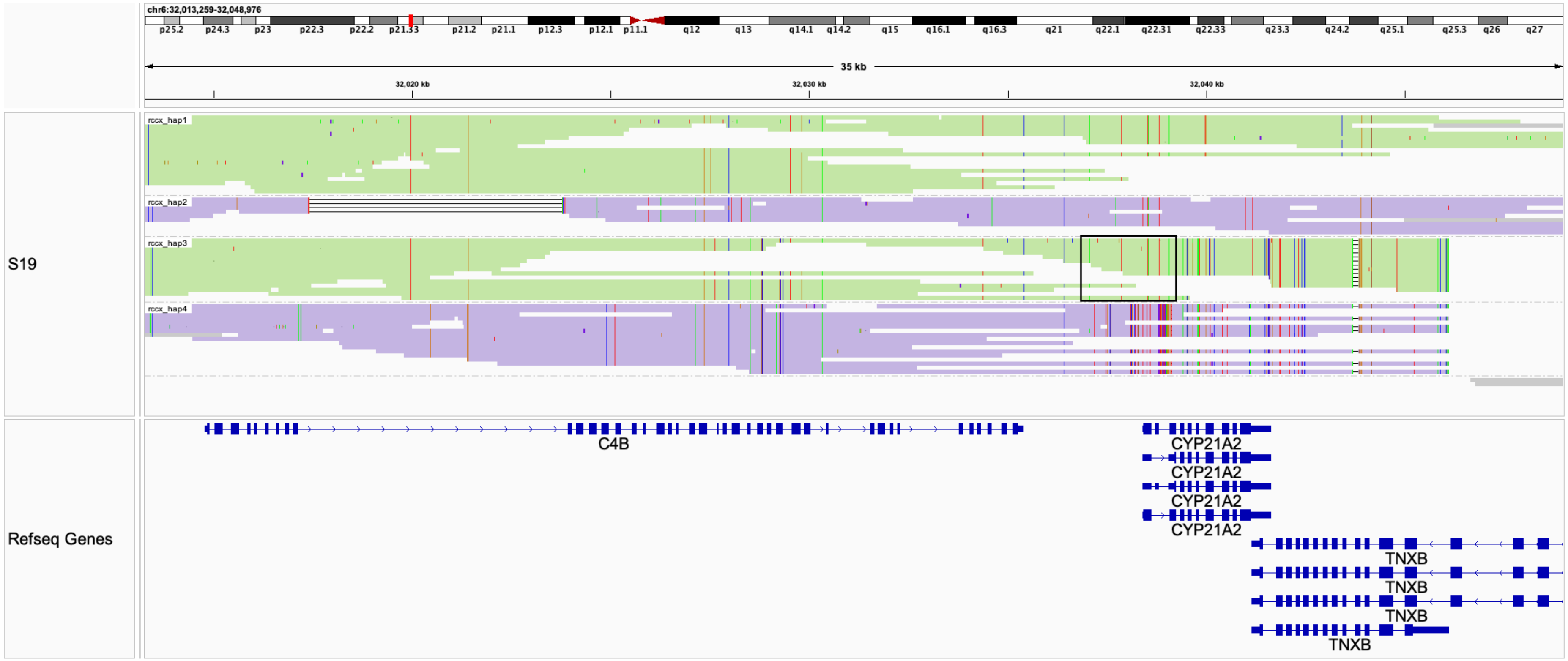


**Figure S4: Sample where MLPA wrongly called a duplication in *CYP21A2*.**

Four copies of the RCCX repeat were phased by Paraphase. Haplotype 1 and 3 (colored green) are on one chromosome and haplotype 2 and 4 (colored purple) are on the other chromosome. Haplotype 1 and 2 extend into the non-homology region downstream and hence are the last copies of the repeat on each allele, encoding *CYP21A2*. Haplotype 3 and 4 are the first copies of the repeat on each allele, encoding the pseudogene *CYP21A1P*. The *CYP21A1P* copy on Haplotype 3 is partially converted to *CYP21A2*-like sequence in the region denoted by the black box. MLPA wrongly detected this gene conversion region in *CYP21A1P* as a duplicated region in *CYP21A2*.

### Supplementary Tables

**Table S1: Overview of all 125 variants in paralogous genes in 86 samples**
External file: Table S1.xlsx

For each of the 125 variants in the 86 samples this table shows the sample ID, variant ID, variant details, variant type, inheritance, original diagnostic detection method, which variant caller the variant identified, the method of detection in lrGS. The paralogous genes that were not supported by Paraphase are marked in the "Remark" column. The 19 samples that were also included in the study by Höps *et al.* (**Methods**)[1], are marked by an asterisk in the "Sample ID" column.

**Table S2: Sequencing characteristics of the 86 samples.**

External file: Table S2.xslx

Columns from left to right: Sample identifier, Gender, Number of HiFi reads, Total HiFi yield, Average HiFi read length, Average HiFi Phred quality score, Average genome sequencing coverage.

**Table S3: Loci supported and not supported by Paraphase.**

| Not supported by Paraphase | Supported by Paraphase |
|---|---|
| *ABCC6* | *CYP21A2* |
| *PKD1* | *IKBKG* |
| *RPGR* | *OTOA* |
| *SHOX* | *PMS2* |
| | *SMN1* |
| | *STRC* |
| | *TNXB** |

* Homology region supported by Paraphase v3.1.2 or earlier; Full gene supported by Paraphase v3.2.0 or later.

### Supplementary Methods

#### Sample selection

Samples (n=86) with one or more known variants in 11 different segmental duplication or other paralogous regions were retrospectively selected from laboratories of Bioscientia in Ingelheim, the Medical University of Innsbruck, the Institute of Clinical Molecular Biology in Kiel and the Radboudumc in Nijmegen. Of note, 31 of the 86 samples contained more than one known genetic variant in the region of interest (range 1-5). This resulted in a total of 125 variants in the 86 samples, which were all individually numbered (**Table S1**). In total, 19 of the 86 samples were also included in a study investigating the performance of lrGS for the detection of difficult-to-detect clinically relevant variants, not only including homologous regions, but also complex rearrangements, repeat expansions, methylation defects, mitochondrial variants, regions of homozygosity and other SVs[1].

This study conforms to the principles of the Helsinki declaration. In addition, this study was approved by center specific Ethics Committees, including the Medical Review Ethics Committee Arnhem-Nijmegen under 2020-7142.

#### PacBio HiFi sequencing and secondary data analysis

Library preparation was performed according to the manufacturer's instructions using Revio V1 chemistry (PacBio, Menlo Park, CA, USA). 7 µg of DNA was sheared on a Megaruptor 3 machine (Diagenode, Liège, Belgium) to a target size of ± 15–18 kb. Libraries were then prepared using SMRTbell prep kit 3.0 (PacBio, Menlo Park, CA, USA), separated on the BluePippin system (Sage Science, Beverly, MA, USA) and size selected to yield >10 kb fragments. Sequencing polymerase was bound to the SMRTbell library using the Revio polymerase kit (PacBio, Menlo Park, CA, USA). Finished libraries were loaded onto a single 25M Revio SMRT cell and sequenced on a Revio instrument (PacBio, Menlo Park, CA, USA), according to the manufacturer's instructions. HiFi reads were aligned to the GRCh38/hg38 reference genome using pbmm2 (v.1.10.0). Structural variants were called using pbsv (v.2.9.0), small variants using DeepVariant (v.1.5.0) and copy number variants were detected using the variation in coverage depth by HifiCNV (v.0.1.6). Finally, Paraphase (v.3.0.0) was run to specifically call variants in paralogs and pseudogenes. Of the 11 different paralogous regions, only seven loci were supported by Paraphase; (https://github.com/PacificBiosciences/paraphase). These regions were selected by Chen *et al.* because they have alignment matches >10kb in size with >99% sequence similarity[2,3]. The remaining four loci were analyzed without Paraphase. All detected variants were annotated using center specific in-house pipelines and publicly available databases.

#### Variant analysis

The data output files were checked for the presence of the known genetic variant(s) in each sample, following the approach described previously[1], except that we replaced the automated step with a manual assessment. In short, the output file associated with the variant class of the variant of interest was used and we specifically searched for the variant of interest. For the samples with variants in Paraphase supported loci, we also searched for the respective variant in the Paraphase output variant call format (vcf) files and Java script object notation (json) file. If the variant was not detected by the standard variant caller or Paraphase, the region of interest was manually inspected in IGV using the general binary alignment map (BAM) file and/or the Paraphase BAM file to look for evidence of the event.

### HiFi Solves EMEA Consortium

Website:

**https://www.pacb.com/hifi-solves/#EMEA-hifisolves**

Co-coordinators:

Sukhvinder Nicklen
PacBio, Menlo Park, CA, USA;


Alexander Hoischen
Department of Human Genetics, Department of Internal Medicine, Radboud University Medical Center, Nijmegen, The Netherlands;


Partners:

1. **Department of Human Genetics, Research Institute for Medical Innovation, Radboud University Medical Center, Nijmegen, the Netherlands**
   Bart van der Sanden; Christian Gilissen; Lisenka Vissers

2. **Institute for Medical Diagnostics GmbH, Bioscientia**
   Christian Betz, Marcel Nebenfuehr, Zafar Yueksel, Hanno Bolz

3. **Department of Biomolecular Medicine, Center for Medical Genetics Ghent University and Ghent University Hospital**
   Brecht Guillemyn, Kim de Leeneer, Elfride De Baere

4. **Division of Computational Biology, Department of Integrative Biomedical Sciences Institute of Infectious Disease and Molecular Medicine, University of Cape Town**
   Mohammed Farahat

5. **Department of Clinical Genetics and Genomics, Karolinska University Hospital, Stockholm, Sweden**
   Kristina Lagerstedt Robinson

6. **Department of Clinical Genetics and Genomics; Department of Molecular Medicine and Surgery, Karolinska University Hospital; Karolinska Institutet**
   Anna Lindstrand

7. **Laboratory of Human Genetics & Therapeutics, Biological and Environmental Sciences and Engineering Division (BESE), King Abdullah University of Science and Technology (KAUST)**
   Bruno Reversade

8. **Division of Clinical Genetics, Department of Laboratory Medicine , Lund University**
Hans Ehrencrona

9. **Institute of Human Genetics, Department of Genetics, Medical University of Innsbruck**
Katharina Herzog, Johannes Zschocke

10. **Clinical Omics and Informatics Unit, Department of Medicine, Neuroscience Institute, University of Cape Town**
Gideon Akuamoah Wiafe, Melissa Nel

11. **Medical and Population Genomics Lab, Department of Translational Medicine, Sidra Medicine**
Sondoss Hassan, Zenab Siddig, Radi Farhad, Mohammadmersad Ghorbani, Rawan Abouelhassan, Melissa Tauro, Younes Mokrab

12. **Institute of Human Genetics, University Hospital Schleswig-Holstein, University of Lübeck and Kiel University, Germany**
Malte Spielmann

13. **Department of Human Genetics, University Hospitals Leuven (UZ Leuven)**
Wouter Bossuyt, Erika Souche, Joris Vermeesch

14. **Institute of Genomics, University of Tartu**
Georgi Hudjasov, Lili Milani

15. **Department of Immunology, Genetics and Pathology, Uppsala University**
Lars Feuk

### References


1. Höps, W., Weiss, M.M., Derks, R., Galbany, J.C., Ouden, A.D., van den Heuvel, S., Timmermans, R., Smits, J., Mokveld, T., Dolzhenko, E., et al. (2025). HiFi long-read genomes for difficult-to-detect, clinically relevant variants. Am J Hum Genet *112*, 450-456. 10.1016/j.ajhg.2024.12.013.
2. Chen, X., Baker, D., Dolzhenko, E., Devaney, J.M., Noya, J., Berlyoung, A.S., Brandon, R., Hruska, K.S., Lochovsky, L., Kruszka, P., et al. (2025). Genome-wide profiling of highly similar paralogous genes using HiFi sequencing. Nature Communications *16*, 2340. 10.1038/s41467-025-57505-2.
3. Chen, X., Harting, J., Farrow, E., Thiffault, I., Kasperaviciute, D., Hoischen, A., Gilissen, C., Pastinen, T., and Eberle, M.A. (2023). Comprehensive SMN1 and SMN2 profiling for spinal muscular atrophy analysis using long-read PacBio HiFi sequencing. Am J Hum Genet *110*, 240-250. 10.1016/j.ajhg.2023.01.001.
